# Post-pandemic ecological reshaping of respiratory pathogen circulation: A six-year FilmArray®-based surveillance study in Tokyo, Japan (2020–2026)

**DOI:** 10.64898/2026.08.28.26360747

**Authors:** Junko S Takeuchi, Masami Kurokawa, Kei Yamamoto, Junko Yamanaka, Eriko Morino, Sakino Takayanagi-Nishisako, Norio Ohmagari, Wataru Sugiura, Moto Kimura

## Abstract

**Background:** The COVID-19 pandemic substantially altered respiratory pathogen circulation worldwide. However, longitudinal analyses of changes in respiratory pathogen ecology across the pandemic and post-pandemic periods remain limited.

**Methods:** We analyzed 19,968 respiratory samples tested with the BioFire® FilmArray® Respiratory Panel at a hospital in Tokyo, Japan, between January 2020 and March 2026. We evaluated temporal changes in pathogen circulation, age-specific epidemiology, co-detection patterns, pairwise pathogen associations, and clinical parameters.

**Results:** At least one respiratory pathogen was detected in 27.8% of tests. Respiratory pathogens resurged asynchronously following the relaxation of COVID-19-related public health measures. Influenza virus circulation remained markedly suppressed until late 2022 before re-emerging in successive large seasonal epidemics, whereas other pathogens, including RSV, human metapneumovirus, and *Mycoplasma pneumoniae*, exhibited distinct resurgence patterns. Pathogen distributions also varied by age. Human rhinovirus/enterovirus remained predominant among young children, whereas SARS-CoV-2 predominated among older adults. Co-detection occurred in 14.0% of positive specimens and was significantly more frequent in younger patients. Pairwise analysis identified both positive and negative pathogen associations; however, the patterns varied across age groups and study periods.

**Conclusions:** Respiratory pathogen circulation changed substantially during the transition from the COVID-19 pandemic to the post-pandemic period, with pathogen-specific, age- and period-dependent patterns. Continued surveillance is warranted to determine how respiratory pathogen circulation will evolve and to inform infection control strategies in the post-pandemic era.

## Introduction

Respiratory tract infections remain a cause of morbidity and mortality worldwide [1]. Multiple respiratory pathogens, including influenza viruses (IFVs), severe acute respiratory syndrome coronavirus 2 (SARS-CoV-2), seasonal coronaviruses (CoVs), respiratory syncytial virus (RSV), adenovirus (AdV), and enterovirus (EV), circulate simultaneously and exhibit distinct epidemiological and seasonal patterns. Rapid multiplex polymerase chain reaction (PCR) assays, such as the BioFire® FilmArray® Respiratory Panel, have enabled comprehensive, real-time detection of respiratory pathogens and improved understanding of respiratory virus ecology in clinical settings [2].

The coronavirus disease 2019 (COVID-19) pandemic dramatically altered the epidemiology of respiratory pathogens worldwide [3]. Non-pharmaceutical interventions, including mask wearing, social distancing, school closures, and travel restrictions, substantially suppressed the transmission not only of SARS-CoV-2 but also of many other respiratory pathogens [3–5]. However, the subsequent relaxation of these measures led to atypical resurgence patterns among multiple respiratory pathogens. Although several studies have described changes in respiratory pathogen circulation during the COVID-19 pandemic [6–8], long-term longitudinal analyses in real-world tertiary-care settings remain limited. Furthermore, comprehensive analyses integrating seasonality, age- and department-specific distributions, co-detection patterns, and associations with clinical parameters remain scarce.

In this study, we retrospectively analyzed respiratory pathogen detection patterns obtained with the BioFire® FilmArray® Respiratory Panel at a tertiary-care hospital from January 2020 to March 2026. We performed comprehensive analyses of temporal and seasonal trends, age- and clinical department-specific distributions, co-detection patterns, and associations with clinical parameters during and after the COVID-19 pandemic to elucidate the post-pandemic ecological reshaping of respiratory pathogens.

## Methods

### Ethics statement

This study was approved by the Ethics Review Committee of the Japan Institute for Health Security (JIHS), Japan (NCGM-004190). All methods were performed in accordance with relevant guidelines and regulations.

### Data collection

This retrospective, single-center study was conducted from January 29, 2020, to March 31, 2026, at the National Center for Global Health and Medicine (NCGM) and the Japan Institute for Health Security (JIHS) in Tokyo, Japan. NCGM is one of four Designated Medical Institutions for Specified Infectious Diseases in Japan and has been involved in the COVID-19 response since the outbreak began. Nasopharyngeal swab samples were tested using the BioFire® FilmArray® Respiratory 2.0 or 2.1 panels (RP2.0 or RP2.1) (bioMérieux, Marcy L’Etoile, France), which rapidly and simultaneously detect a wide range of respiratory pathogens. The RP2.0 panel detects 17 viral, 2 bacterial, and 2 atypical bacterial pathogens (adenovirus [AdV]; coronaviruses 229E, HKU1, OC43, and NL63 [CoV]; human metapneumovirus [hMPV]; human rhinovirus/enterovirus [HRV/EV]; influenza A, A/H1, A/H1-2009, A/H3, and B viruses [IFV]; human parainfluenza viruses 1, 2, 3, and 4 [HPIV]; respiratory syncytial virus [RSV]; *Bordetella parapertussis* [*B. parapertussis*]; *Bordetella pertussis* [*B. pertussis*]; *Chlamydia pneumoniae* [*C. pneumoniae*]; and *Mycoplasma pneumoniae* [*M. pneumoniae*]), and the RP2.1 panel additionally detects SARS-CoV-2.

### Data management

This study included all diagnostic data obtained at NCGM using the RP2.0 or RP2.1 panels during the study period. Tests from the same patient were otherwise treated as independent observations.

However, when multiple results were obtained within 7 days, these tests were considered part of the same testing episode, and only the most recent result was retained to avoid duplication due to repeat testing or interdepartmental transfers. For each test, the following data were collected: test date, detected pathogens, sex, age, body temperature, C-reactive protein (CRP) level, and the clinical department that ordered the test.

In this study, the positivity rate was defined as the proportion of tests that were positive for a given pathogen among all tests, unless otherwise specified. For analyses based on positive detections, however, it was calculated as the proportion of positive detections for each pathogen among all positive detections. Co-detection was defined as the detection of two or more pathogens in a single test. Seasons were defined as spring (March to May), summer (June to August), autumn (September to November), and winter (December to February). Epidemic years were defined as extending from December to November; for example, epidemic year 2022 corresponded to the period from December 2021 through November 2022. The study period was stratified into five categories reflecting major changes in the epidemiological and testing context: pre-pandemic (through July 2020), pandemic 1 (August 2020–December 2021), pandemic 2 (January 2022–November 2022), pandemic 3 (December 2022–May 7, 2023), and post-pandemic (from May 8, 2023, onward). Four age groups were defined: children (0–5 years), school-age children (6–17 years), adults (18–59 years), and older adults (≥60 years) (see Supplementary Method 1 for more details).

### Clinical characteristics

The distributions of patient body temperature and CRP levels were evaluated for each pathogen group. To exclude the influence of co-detection, only test records in which a single pathogen was detected were included. Furthermore, only records collected during the post-pandemic period, when symptomatic cases were more prevalent, were included.

### SARS-CoV-2 variants

SARS-CoV-2 variants before December 2024 were primarily identified by Sanger sequencing, as previously reported [9]. From January 2025 onward, variant data were additionally obtained using amplicon-based next-generation sequencing (NGS) (see Supplementary Method 2 for more details).

### Descriptive and statistical analysis

All descriptive and statistical analyses were performed using R version 4.3.1 (R Foundation for Statistical Computing, Vienna, Austria). The frequencies of single- and multiple-pathogen detections were summarized using an intersection-based approach implemented with the ComplexUpset package (v1.3.5) [10], capturing all observed pathogen combinations. Pairwise co-detection patterns were also visualized using heatmaps, with diagonal cells representing single-pathogen detection frequencies. The co-detection heatmap analyses were restricted to samples with at least one detected pathogen. Pairwise associations between pathogens were evaluated using 2×2 contingency tables, as previously described [11] with minor modifications, using all tested samples, including those with no pathogen detected (see Supplementary Method 3 for more details).

Differences in patient age according to the number of detected pathogens were evaluated using the Jonckheere–Terpstra trend test in the clinfun package (v1.1.5).

Body temperature and CRP levels were compared across multiple pathogen groups using the Kruskal-Wallis test; post hoc comparisons, performed only if p < 0.05, were conducted using Dunn’s test with Benjamini–Hochberg false discovery rate (FDR) correction. To evaluate whether differences in body temperature and CRP levels were independent of age, a multivariate linear regression model was fitted using the *lm()* function. SARS-CoV-2 and *M. pneumoniae* were selected as the reference categories for the body temperature and CRP analyses, respectively.

## Results

### Study population

Between January 29, 2020, and March 31, 2026, a total of 20,281 BioFire® FilmArray® Respiratory Panel tests were performed on 16,014 unique patients. Most patients (81.5%) underwent the tests once. Repeated testing was observed in 21.8%, 13.7%, 15.9%, and 14.6% of children, school-age children, adults, and older adults, respectively. The median interval between testing episodes was 92.5 days (interquartile range [IQR]: 32–239 days). After retaining only the most recent result for patients who underwent multiple tests within 7 days, 19,968 test results remained for further analysis. The median patient age at testing was 51 years (IQR: 23–77). Males accounted for 52.6% of test records. During the study period, the population eligible for testing changed for several reasons, as described in Supplementary Figure 1A and B and Supplementary Method 1, and the age distribution differed across periods (Supplementary Figure 1C, Supplementary Table 1). Accordingly, the study period was divided into five phases.

### Overall detection rate

Among the 19,968 tests analyzed, at least one respiratory pathogen was detected in 27.8% (n=5,560) of samples, including viral pathogens in 27.2% (n=5,429) and bacterial and atypical bacterial pathogens in 1.0% (n=204) (Table 1). HRV/EV was the most frequently detected pathogen (9.6%, n=1,918), followed by SARS-CoV-2 (8.3%, n=1,662), RSV (3.3%, n=652), HPIV (2.9%, n=576), and IFV (2.1%, n=426) (Supplementary Table 2). Throughout the pandemic phases, SARS- CoV-2 was the most frequently detected pathogen, followed by HRV/EV. In contrast, during the post-pandemic period, which was characterized by selective testing of symptomatic patients, HRV/EV became the predominant pathogen (20.9%, n=1,074), followed by SARS-CoV-2 (9.4%, n=484), HPIV (7.3%, n=374), RSV (6.9%, n=357), and IFV (6.8%, n=352).

**Table 1.** Positivity rates of viral and bacterial pathogens.

|  | Tested | All |  | Virus |  | Bacterial and atypical bacterial pathogens |  |
| --- | --- | --- | --- | --- | --- | --- | --- |
|  |  | Positive | Proportion | Positive | Proportion | Positive | Proportion |
| **All** | 19,968 | 5,560 | 27.8% | 5,429 | 27.2% | 204 | 1.0% |
| **Sex** |  |  |  |  |  |  |  |
| Female | 9,462 | 2,665 | 28.2% | 2,601 | 27.5% | 87 | 0.9% |
| Male | 10,505 | 2,894 | 27.5% | 2,827 | 26.9% | 117 | 1.1% |
| Unknown | 1 | 1 | - | 1 | - | - | - |
| **Age group** |  |  |  |  |  |  |  |
| Children (0-5) | 2,930 | 2,175 | 74.2% | 2,138 | 73.0% | 78 | 2.7% |
| School-age children (6-17) | 1,197 | 588 | 49.1% | 531 | 44.4% | 82 | 6.9% |
| Adults (18-59) | 7,137 | 2,006 | 28.1% | 1,973 | 27.6% | 39 | 0.5% |
| Older adults (60+) | 8,700 | 790 | 9.1% | 786 | 9.0% | 5 | 0.1% |
| Unknown | 4 | 1 | - | 1 | - | - | - |
| **Study period** |  |  |  |  |  |  |  |
| Pre-pandemic (–Jul 2020) | 193 | 38 | 19.7% | 34 | 17.6% | 4 | 2.1% |
| Pandemic 1 (Aug 2020–Dec 2021) | 10,754 | 1,006 | 9.4% | 1,004 | 9.3% | 2 | 0.0% |
| Pandemic 2 (Jan 2022–Nov 2022) | 2,125 | 1,066 | 50.2% | 1,066 | 50.2% | - | - |
| Pandemic 3 (Dec 2022–May 7, 2023) | 1,748 | 595 | 34.0% | 594 | 34.0% | 1 | 0.1% |
| Post-pandemic (May 8, 2023–) | 5,148 | 2,855 | 55.5% | 2,731 | 53.0% | 197 | 3.8% |

Positivity rates were higher during the post-pandemic period than during the pandemic period (Table 1 and Supplementary Table 1). This difference likely reflects changes in testing strategies: universal screening was widely implemented during the pandemic, whereas post-pandemic testing was primarily limited to symptomatic patients. Positivity rates also varied across epidemic periods and by patient age. The results for each category are summarized below.

### Detection trends and seasonality of respiratory pathogens

Distinct seasonal patterns and disruptions of conventional seasonality were observed among respiratory pathogens (Figure 1 and Supplementary Figure 2). The detection frequencies of several respiratory pathogens were markedly reduced during the COVID-19 pandemic. Soon after the COVID-19-related public health measures were relaxed during the spring and summer of 2021, CoV-NL63, RSV, and HPIV-3 showed increased detection rates.

**Figure 1.**
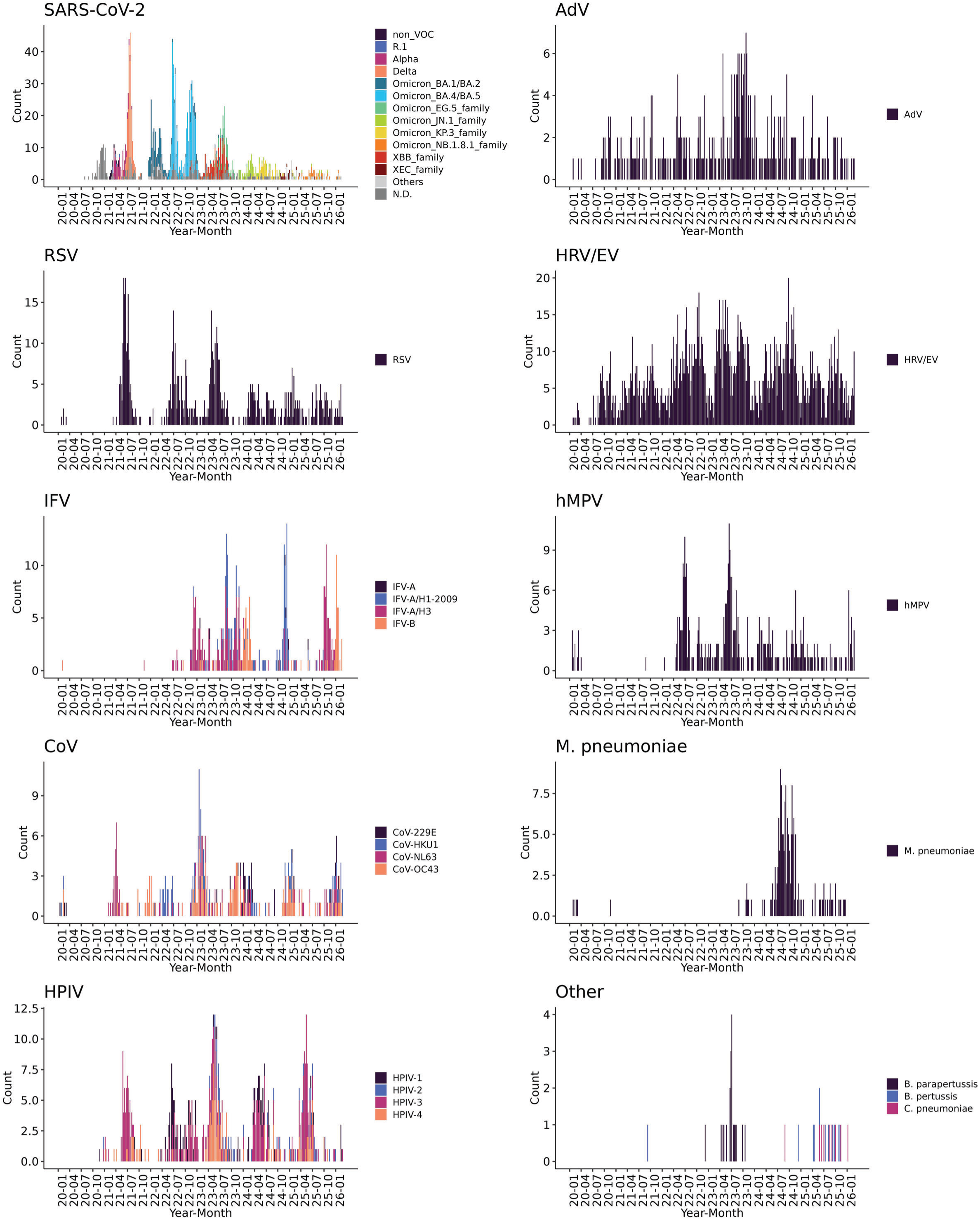
Temporal distribution of respiratory pathogens detected using the BioFire® FilmArray® Respiratory Panel from January 2020 to March 2026. Weekly counts of detected pathogens are shown for each respiratory pathogen. The y-axis shows the number of positive samples, with colors indicating individual species, lineages, or subtypes, as appropriate. The x-axis shows the year and month of sample collection. N.D., not determined.

IFV detection was particularly limited during pandemic period. In early 2023, as SARS-CoV-2 Omicron variants circulated, IFV A (H3) predominated; later that year, detection of IFV A (H1) pdm09 increased. Following substantial detection of IFV B in early 2024, IFV A (H1) pdm09 predominated during the winter of the 2024–2025 influenza season. In late 2025, IFV A (H3) increased again, followed by a peak in IFV-B detection in February 2026.

Increased detection of hMPV was observed during the summers of 2022 and 2023, whereas *M. pneumoniae* increased in the summer and autumn of 2024. In contrast, HRV/EV were detected continuously throughout the study period, although its detection frequency decreased during the COVID-19 pandemic.

### Age-specific positivity rates

Children exhibited a wide diversity of respiratory pathogens throughout the study period. In contrast, pathogen diversity decreased with age, particularly during the pandemic, when detections were largely dominated by SARS-CoV-2 (Figure 2). Based on the proportion of positive detection, in the 2021 epidemic year, SARS-CoV-2 accounted for 65% and 78.5% of total positive detections among adults and older adults, respectively. In contrast, HRV/EV was the predominant pathogen among children and school-age children (Supplementary Figure 3). During the 2022 epidemic year, when SARS-CoV-2 Omicron variants became predominant, SARS-CoV-2 was the most frequently detected pathogen among adults, older adults, and school-age children. However, HRV/EV remained the predominant pathogen among children. Since the 2023 epidemic year, the proportion of SARS- CoV-2 detections has gradually declined across all age groups. Nevertheless, SARS-CoV-2 remained the predominant pathogen among older adults through the 2025 epidemic year.

**Figure 2.**
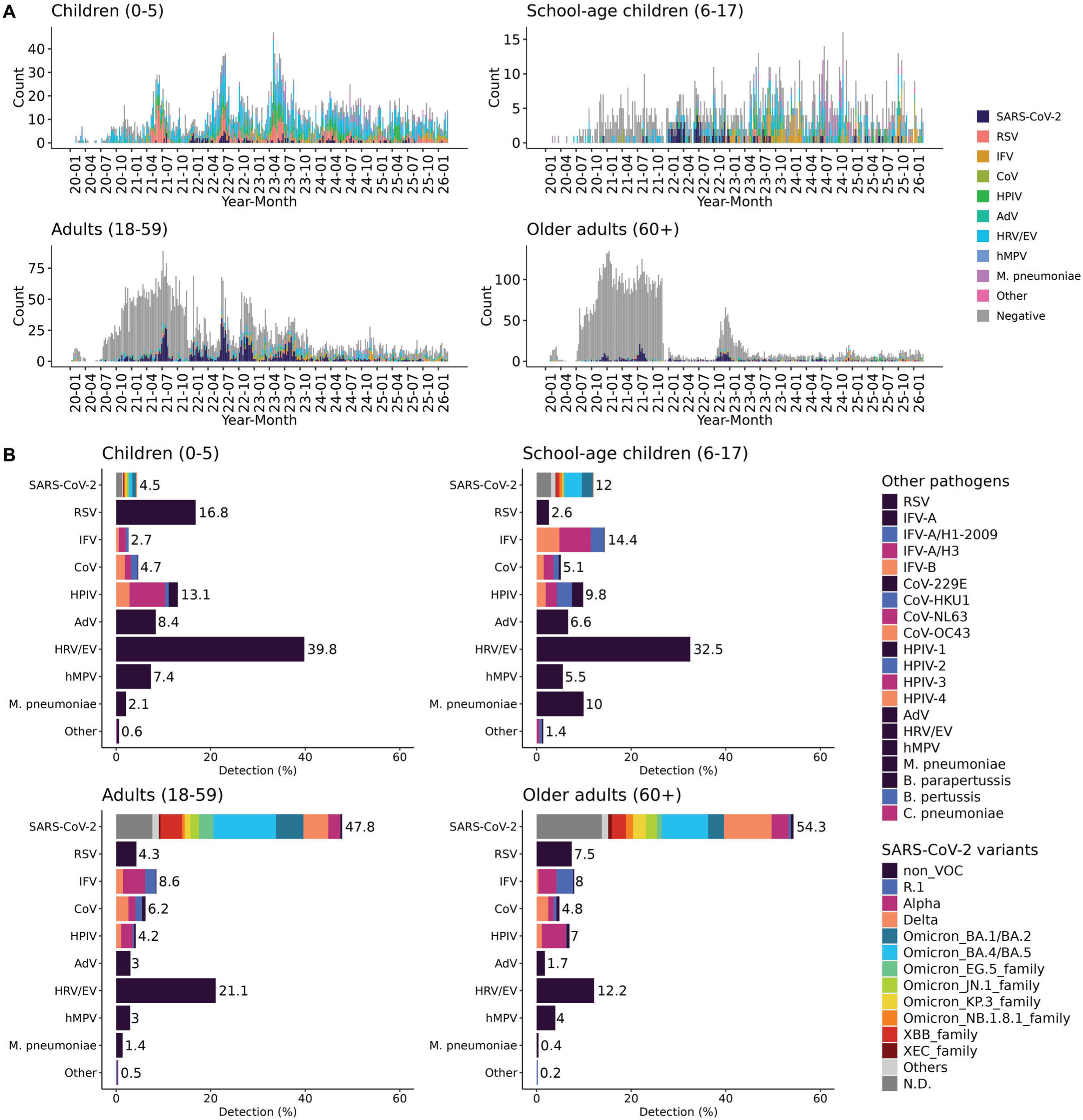
Distribution of respiratory pathogens by age group. **A.** Weekly counts of all tested samples are shown for four age groups (0–5, 6–17, 18–59, and ≥60 years). The y-axis shows the number of test results, with multiple pathogen detections from a single sample counted separately; colors indicate the detected respiratory pathogens, and gray indicates samples in which no pathogen was detected. The x-axis shows the year and month of sample collection. **B.** The overall composition of respiratory pathogens is shown by age group. Bar lengths and the numbers shown beside them indicate the proportion of each respiratory pathogen, calculated as the number of positive detections for each pathogen divided by the total number of positive detections within each age group. Colors indicate individual species, lineages, or subtypes, as appropriate. N.D., not determined.

### Department-specific positivity rates

Testing was requested by 39 clinical departments, including Emergency (n=7,964), Pediatrics (n=3,690), Infectious Diseases (n=3,059), and Respiratory Medicine (n=887). Pathogen detection rates also varied markedly across clinical departments. Among departments that ordered more than 500 tests and had a positivity rate exceeding 10%, Pediatrics had the highest positivity rate (68.3%; 2,521/3,690), consistent with the higher detection rates observed in children. This was followed by Infectious Diseases (46.4%; 1,419/3,059), General Medicine & Infectious Diseases (34.0%; 181/533), Respiratory Medicine (22.3%; 198/887), Internal Medicine (14.8%; 77/522), and Emergency (11.1%; 882/7,964) (Supplementary Figure 4).

### Co-detection patterns

Among the 5,560 positive tests, single-pathogen detections accounted for 86.0% (n=4,780), whereas co-detections accounted for 14.0% (n=780). Among the co-detected cases, dual-pathogen detections were most common (n=649; 11.7%), followed by triple (n=103; 1.8%), quadruple (n=21; 0.38%), and quintuple detections (n=7; 0.13%) (Supplementary Figure 5). Patient age decreased significantly as the number of co-detected pathogens increased (Jonckheere–Terpstra trend test, P < 0.001).

ComplexUpset analysis demonstrated heterogeneous co-detection patterns among respirator pathogens (Figure 3A). The most common co-detection pattern was HRV/EV plus RSV (n=89). Notably, all eight of the most frequent co-detection patterns involved HRV/EV in combination with other pathogens, including AdV (n=87), HPIV-3 (n=62), hMPV (n=43), *M. pneumoniae* (n=32), SARS-CoV-2 (n=31), HPIV-4 (n=26), and HPIV-1 (n=19).

**Figure 3.**
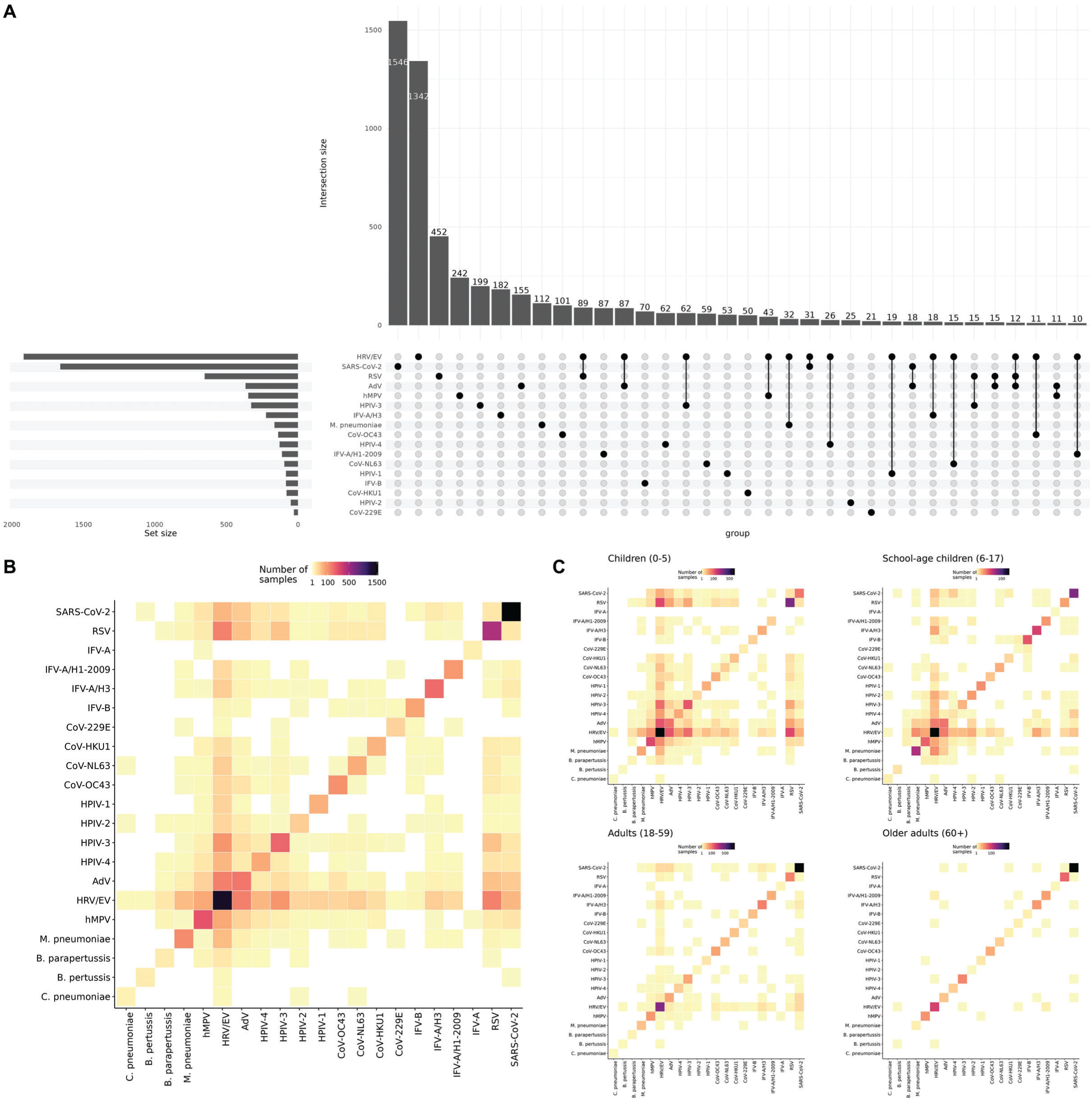
Co-detection patterns among respiratory pathogens. The ComplexUpset plot visualizes the distribution of single- and multiple-pathogen detections observed in ≥10 samples. Dots and connecting lines show the pathogen composition of each intersection **(A)**. The heatmap illustrates pairwise co-detection patterns among respiratory pathogens **(B)**, and the heatmaps show these patterns stratified by age group **(C)**. The analyses were restricted to samples with at least one detected pathogen. Each cell represents the number of samples in which the corresponding pathogen pair was detected, with diagonal cells representing single-pathogen detection frequencies. Color intensity indicates the number of samples, with lower and higher values represented by yellow and dark purple, respectively.

Heatmap visualization highlighted both the predominance of single-pathogen detections and distinct co-detection relationships among pathogen pairs (Figure 3B). Age-stratified heatmaps showed that co-detection patterns were most extensive among children and gradually diminished with increasing age (Figure 3C). Pairwise analysis identified both positive and negative associations after FDR correction (Figure 4A). However, the strength and direction of pathogen associations varied across age groups (Figure 4B). Furthermore, stratification by both age group and study period revealed additional variation in pathogen co-detection patterns (Supplementary Figure 6) and associations (Supplementary Figure 7). Although co-detections were more frequent during the post- pandemic period, the specific patterns of association differed across age groups and study periods.

**Figure 4.**
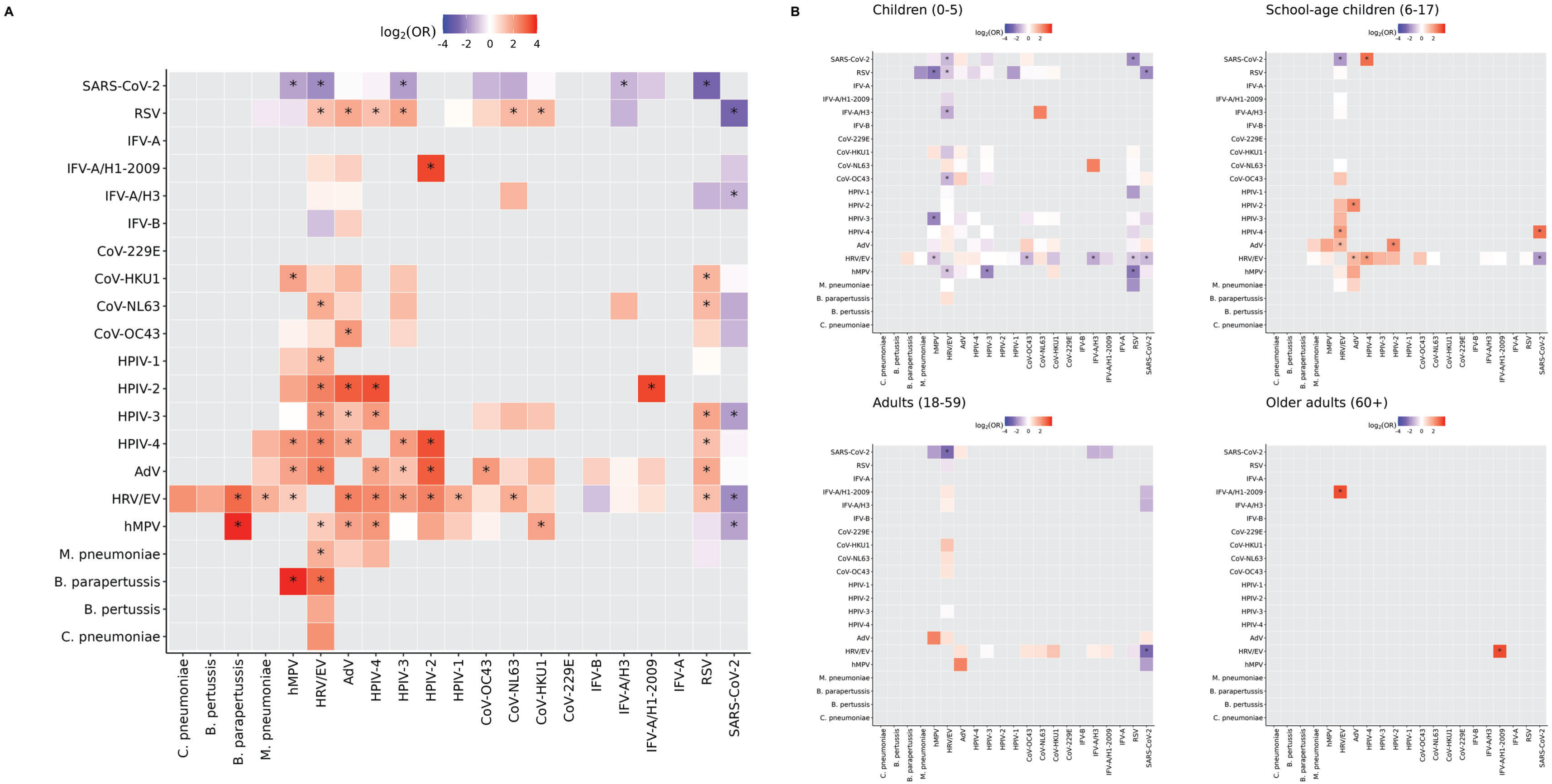
Association patterns among respiratory pathogens. The heatmap shows statistical pairwise associations between pathogen pairs based on log2-transformed odds ratios (ORs) **(A)**, and the heatmaps show these patterns stratified by age group **(B)**. Red and blue indicate positive and negative associations, respectively, and color intensity represents the strength of the association. Significant associations after false discovery rate (FDR) correction are indicated by an asterisk.

### Body temperature and C-reactive protein (CRP) levels

Finally, clinical characteristics were evaluated among patients with single-pathogen detection during the post-pandemic period. Body temperature was lower in SARS-CoV-2 group than in several other pathogen groups, and these differences remained significant after adjustment for age. CRP levels tended to be higher in patients with *M. pneumoniae* and other bacterial and atypical bacterial infections (see Supplementary Figure 8 for more details).

## Discussion

In this large longitudinal study of ∼20,000 multiplex respiratory pathogen PCR tests conducted at a tertiary-care hospital in Tokyo, Japan, we demonstrated changes in respiratory pathogen ecology across the COVID-19 pandemic and into the post-pandemic period. Our findings yielded three main observations. First, respiratory pathogen circulation resurged asynchronously following the relaxation of non-pharmaceutical interventions. Second, respiratory pathogen distributions varied markedly by patient age. Third, co-detection analyses revealed distinct patterns of respiratory pathogen associations.

Consistent with findings from global surveillance studies [12], respiratory pathogen detection at our hospital resumed asynchronously across pathogens following the relaxation of COVID-19- related public health measures. The detection patterns of several pathogens, including SARS-CoV-2, RSV, IFV, and *M. pneumoniae*, generally corresponded to nationwide epidemiological trends of their associated diseases reported in the Infectious Disease Weekly Report (IDWR) [13], published by the National Institute of Infectious Diseases (NIID) in Japan (Supplementary Figure 9). The timing of resurgence varied among pathogens. CoVs, RSV, and HPIVs resurged soon after these measures were relaxed. Notably, RSV resurged during the spring and summer of 2021, coinciding with the spread of the SARS-CoV-2 Delta variant and the period surrounding the Tokyo Olympic and Paralympic Games. During this period, the pediatric ward at our hospital was temporarily closed between March and July 2021 in response to the spread of respiratory infections.

IFVs remained markedly suppressed until late 2022 before re-emerging in repeated large seasonal epidemics. Although IFVs typically circulate in Japan from December to March [14], their post- pandemic circulation showed substantial temporal variation, with successive predominance of different subtypes and repeated large epidemic waves. Remarkably, during the 2024–2025 season, Japan recorded its highest weekly number of influenza cases since national surveillance began in 1999 [15]. Furthermore, the 2025–2026 season saw the first occurrence of two national alerts within a single influenza season.

hMPV exhibited atypical summer activity, although this virus typically peaks in spring [16]. AdV detection increased in late 2023 and coincided with the pharyngoconjunctival fever epidemic reported during the same period. In addition, a delayed resurgence of *M. pneumoniae* [17] was observed in 2024, corresponding to the second-largest epidemic reported in Japan during the preceding decade.

In contrast, HRV/EV were detected continuously throughout the study period. Epidemics of herpangina and HFMD (hand, foot, and mouth disease), which are caused by EV, were reported during the study period. However, because the FilmArray® Respiratory Panel does not differentiate HRV from EV, HRV/EV-positive results may not reliably reflect EV-associated diseases (Supplementary Figure 9). Furthermore, the nationwide resurgence of pertussis in 2025, which reached the highest number of cases since nationwide case-based surveillance began in 2018 (Supplementary Figure 9), was not clearly reflected in our FilmArray®-based surveillance. During this period, FilmArray® testing at our hospital mainly targeted referred or hospitalized patients with fever or respiratory symptoms, whereas many patients with pertussis presented with prolonged paroxysmal cough without fever and were diagnosed as outpatients using pertussis-specific tests such as LAMP.

These heterogeneous post-pandemic resurgence patterns were observed across a broader range of respiratory pathogens. This observation may be partly explained by the “immunity debt” hypothesis, which posits that reduced exposure to respiratory pathogens during periods of restricted social contact may have increased the proportion of susceptible individuals at the population level [18,19]. In addition, several pathogen-specific characteristics, including differences in the durability of protective immunity, pre-existing community circulation, pathogen interference, and transmissibility, may explain these asynchronous and distinct resurgence patterns [12,20]. Thus, respiratory pathogen circulation is likely shaped by multiple factors and has not simply returned to the pre-pandemic state. Whether these patterns will return to typical circulation or stabilize into a new normal remains uncertain [21], underscoring the importance of continued comprehensive monitoring of respiratory pathogens. Automated surveillance systems based on multiplex PCR diagnostic data, such as *Syndromic Trends*, have been developed for real-time analysis of circulating respiratory pathogens and the detection of new outbreaks [2].

Age-dependent differences in respiratory pathogen epidemiology were observed throughout the study. HRV/EV remained predominant among young children, whereas SARS-CoV-2 consistently accounted for the largest proportion of detections among older adults. These findings likely reflect differences in host immunity, exposure patterns, social behavior, and healthcare utilization across age groups. Despite post-pandemic changes in pathogen circulation, age-specific distributions remained consistent, suggesting that host age remains a key determinant of respiratory pathogen ecology and should be considered when interpreting multiplex PCR results.

Co-detection was identified in 14% of positive specimens and occurred predominantly in younger children. HRV/EV was also the most frequently detected and co-detected pathogen throughout the study period. However, previous studies have demonstrated prolonged detection of HRV/EV RNA for weeks, particularly in young children and immunocompromised individuals [22]. Therefore, some HRV/EV-positive results in this study may represent prolonged viral shedding or residual nucleic acid detection rather than active infection. Furthermore, the FilmArray® Respiratory Panel cannot distinguish EV from HRV and does not provide quantitative viral load values. A previous study at a hospital in Tokyo found that HRV accounted for most FilmArray® HRV/EV-positive specimens (103/115, 89.6%) [23]. This finding is consistent with our observation that HRV/EV was frequently detected outside epidemics of EV-associated diseases, whereas detection of several respiratory pathogens corresponded to peaks in the associated diseases reported by NIID (Supplementary Figure 9). This limitation is clinically relevant because repeated HRV/EV detection is common in routine clinical practice and complicates interpretation of the results. Molecular typing based on capsid-coding sequences (e.g., VP1) [24] may help distinguish persistent viral detection from reinfection with a different HRV/EV type. Therefore, co-detection identified by multiplex PCR should be interpreted in the clinical context and does not necessarily represent simultaneous active infections.

Pairwise association analysis identified several significant positive and negative associations among pathogens. Interestingly, all significant negative associations in the overall analysis involved SARS-CoV-2, consistent with observations from previous epidemiological [25] and experimental [26,27] studies. Experimental studies have demonstrated that infection with one respiratory virus can induce interferon-mediated antiviral responses that suppress subsequent SARS-CoV-2 replication.

However, interpreting these associations at the clinical level requires caution. In our age- and period- stratified analyses, the strength and direction of pathogen associations differed substantially across age groups and study periods. These findings indicate that overall associations may reflect a combination of age- and period-dependent pathogen distributions, as shown in Figure 4 and Supplementary Figure 7, and could also be influenced by shared seasonality and potential pathogen- pathogen interactions. These observations emphasize the complexity of interpreting surveillance data, and further studies integrating epidemiological data with experimental virological evidence are needed.

Finally, although CRP levels tended to be higher in patients with bacterial and atypical bacterial infections, values overlapped across pathogen groups. Likewise, patients with SARS-CoV-2 infection had lower body temperatures than those with RSV or hMPV. Because this analysis included only post-pandemic cases, almost all SARS-CoV-2 infections were attributable to Omicron lineages, which may partly explain the reduced clinical severity compared with earlier variants, including Delta [28]. Overall, the considerable overlap in CRP levels and body temperature across pathogen groups suggests that these routinely available clinical parameters alone are insufficient to identify the causative pathogen [29].

This study has several strengths, including its large sample size, observation period of more than 6 years, consistent use of uniform diagnostic data, and comprehensive analyses integrating epidemiological and clinical characteristics. However, it also has several limitations. First, our data should be interpreted with caution because the testing population changed throughout the study period. During the pandemic, FilmArray® testing was frequently used for universal screening of hospitalized or preoperative patients, whereas post-pandemic testing was largely restricted to symptomatic individuals. Second, patients with a known history of exposure to specific pathogens or with clinical features suggestive of a particular pathogen were more likely to undergo pathogen- specific testing rather than FilmArray® testing. Thus, pathogens that could be clinically suspected or specifically tested for may have been underestimated in our study. Third, this was a single-center retrospective study conducted at a tertiary-care hospital, which served as a major center for COVID- 19 patient care during the pandemic, limiting the generalizability of the results to broader populations and other settings. Finally, detailed clinical outcomes, such as pneumonia and disease severity, were not evaluated, and clinical parameters such as body temperature and CRP levels were limited by missing data.

## Conclusion

As multiple respiratory pathogens continue to co-circulate following the COVID-19 pandemic, multiplex PCR surveillance can provide valuable information for understanding the evolving ecological reshaping of respiratory pathogens, supporting the interpretation of multiplex PCR results in clinical practice, and informing infection control strategies in the post-pandemic era.

## Author contributions

M. Kimura and M. Kurokawa conceived the study. M. Kurokawa performed diagnostic testing and managed study data. J.S.T. designed and performed epidemiological and statistical analyses, prepared the figures, and wrote the original draft of the manuscript. J.S.T. and S.T-N. conducted the SARS-CoV-2 variant analysis. K.Y., J.Y., E.M., N.O., and W.S. contributed to the clinical interpretation of the findings and critically reviewed the manuscript. M. Kimura supervised the project and acquired funding. All authors reviewed and approved the final version of the manuscript.

## Acknowledgements

We thank the staff of Department of Laboratory Testing and all clinical departments at the National Center for Global Health and Medicine (NCGM) for their contributions to this study. We thank Kento Fukano, Azusa Kamikawa, Emiko Hatano, Ryoko Tamura, and Yurika Tanaka for their technical support.

## Data availability statement

The data that support the findings of this study, excluding those subjects to privacy or ethical restrictions, are available from the corresponding authors upon reasonable request.

## Abbreviations

SARS-CoV-2: severe acute respiratory syndrome coronavirus 2
COVID-19: coronavirus disease 2019
AdV: adenovirus
CoV: coronavirus
hMPV: human metapneumovirus
HRV/EV: human rhinovirus/enterovirus
IFV: influenza virus
HPIV: human parainfluenza virus
RSV: respiratory syncytial virus
*B. parapertussis*: *Bordetella parapertussis*
*B. pertussis*: *Bordetella pertussis*
C. pneumoniae: Chlamydia pneumoniae
MP and *M. pneumoniae*: *Mycoplasma pneumoniae*
CRP: C-reactive protein.

