## Supplementary Information for "Post-pandemic ecological reshaping of respiratory pathogen circulation: A six-year FilmArray®-based surveillance study in Tokyo, Japan (2020–2026)"

This PDF file includes:

- Supplementary Methods 1–3
- Supplementary Figures 1–9
- Supplementary Tables 1–2

### Supplementary methods

#### Supplementary Method 1: Rationale for the Definition of Study Periods and Epidemiological Context in Japan

In Japan, the BioFire® FilmArray® RP2.1, which detects SARS-CoV-2 in addition to the targets included in RP2.0, was approved as an *in vitro* diagnostic (IVD) by the Pharmaceuticals and Medical Devices Agency (PMDA) on June 2, 2020, and was subsequently covered by the national health insurance system on July 22, 2020. Therefore, from August 2020 onward, most tests were performed using RP2.1 rather than RP2.0.

During the COVID-19 pandemic, universal SARS-CoV-2 screening of hospitalized and preoperative patients was implemented across multiple departments, regardless of symptoms. FilmArray® testing was requested by 39 clinical departments during the study period. Because the supply of SARS-CoV-2 IVD reagents was unstable, testing strategies changed over time. On May 8, 2023, the Japanese government reclassified COVID-19 as a “Class 5 infectious disease” under the Infectious Diseases Control Law, equivalent to seasonal influenza. Given these circumstances, the study period was divided into five phases: (1) through July 2020, pre-pandemic; (2) August 2020 to December 2021, when FilmArray® was predominantly used; (3) January 2022 to November 2022, when multiple IVD platforms were used concurrently; (4) December 2022 to May 7, 2023, when FilmArray® testing was performed selectively, including in the Emergency Department, in response to the first post-pandemic IFV resurgence and continued SARS-CoV-2 circulation; and (5) after May 8, 2023, when universal testing was primarily replaced by antigen testing and FilmArray® testing was used selectively for patients requiring further evaluation, particularly those presenting with fever and/or respiratory symptoms. Detection rates varied among phases, reflecting differences in testing strategies and patient populations.

#### Supplementary Method 2: Amplicon-based next-generation sequencing for SARS-CoV-2 variant analysis

Nucleic acid (60 µL) was extracted from 200 µL of residual nasopharyngeal swab samples using a KingFisher APEX System (Thermo Fisher Scientific, Waltham, MA, USA) and the MagMAX Viral/Pathogen NA Nucleic Acid Isolation Kit (Thermo Fisher Scientific). Subsequently, cDNA synthesis, target amplification, and library preparation were performed according to the Illumina COVIDSeq Test Reference Guide (Illumina Inc., San Diego, CA, USA) using ARTIC V5.4.2 primers. SARS-CoV-2 genome sequencing was performed using the Illumina iSeq 100 or NextSeq 2000 system. Raw FASTQ files underwent quality control using FastQC (v0.11.7) and adapter and quality trimming using TrimGalore (v0.6.4). The reads were then aligned to the reference genome (Wuhan-Hu-1; MN908947.3) using BWA-MEM (v0.7.17-r1188). The resulting alignments were sorted and indexed using SAMtools (v1.17), and primer sequences were trimmed using iVar trim (v1.4.4), retaining reads without primer matches. Consensus sequences were generated using iVar consensus (v1.4.4), with a minimum depth threshold of 10 and a minimum base quality of 20. Variant calling was additionally performed using bcftools (v1.17). Lineages were assigned using pangolin (v4.3.1) with the corresponding pangolin-data version.

##### Supplementary Method 3: Pairwise associations between pathogens

Pairwise associations between pathogens were evaluated using 2×2 contingency tables, as previously described [11] with minor modifications, using all tested samples, including those with no pathogen detected. Briefly, odds ratios (ORs) and 95% confidence intervals (CIs) were calculated for each pathogen pair. When any expected cell frequency was <5, Fisher's exact test was used; otherwise, the chi-squared test of independence was used. If any value in a contingency table was 0, the Haldane–Anscombe correction was applied to calculate the ORs by adding 0.5 to all cells to accommodate possible zero counts. Pathogen pairs with co-detection counts <3 were excluded from the association analysis. Corresponding P values were adjusted using the Benjamini–Hochberg false discovery rate (FDR) method, and adjusted P values < 0.05 were considered statistically significant. Associations were visualized as a heatmap of log2-transformed ORs, with significant associations indicated by an asterisk.

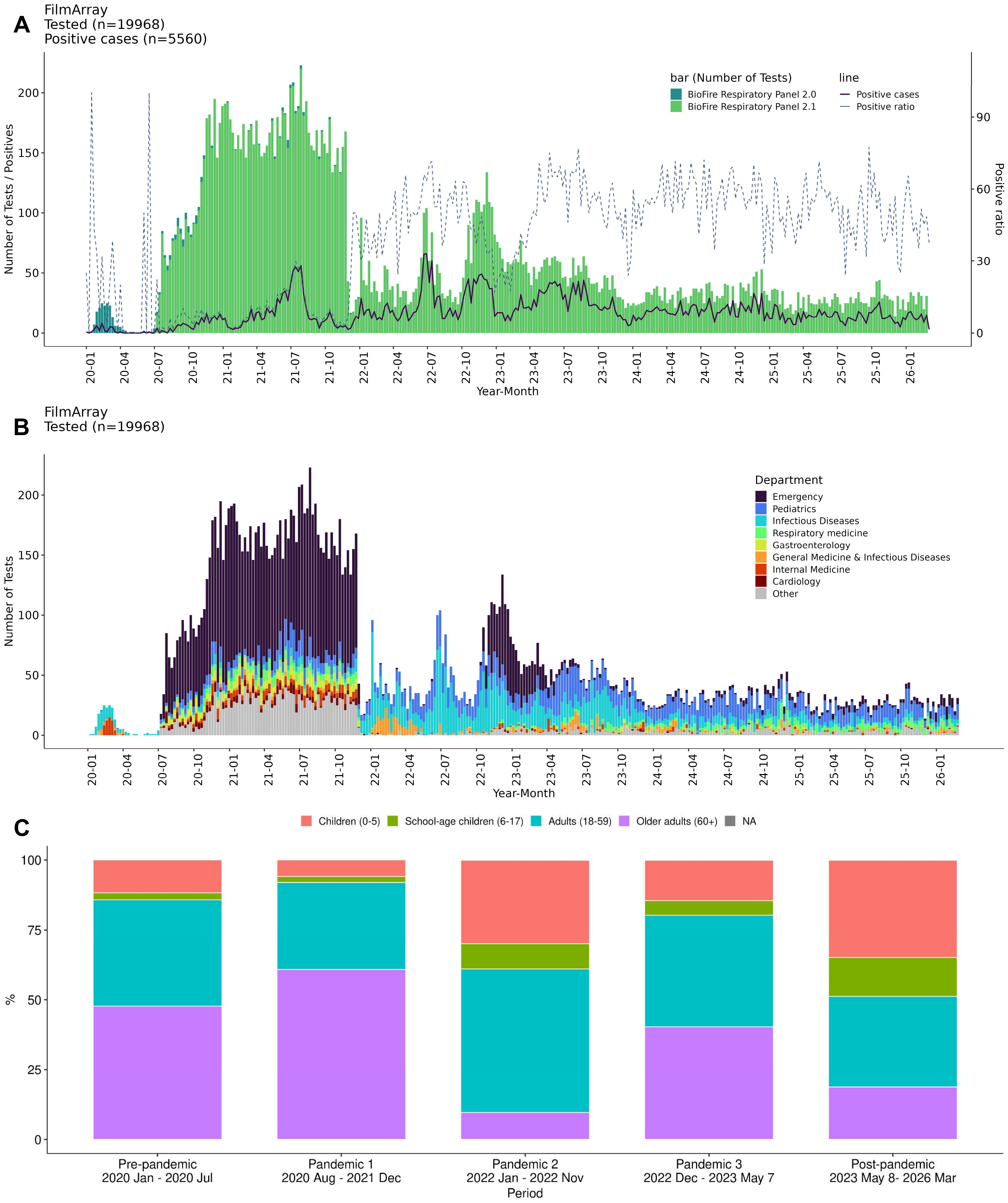


**Supplementary Figure 1. Epidemiological distribution of respiratory pathogens and the study population during the study period. A.** Weekly numbers of tests (bars), positive samples (solid line), and the positivity rate (dashed line) are shown, with colors indicating panel versions. **B.** Weekly numbers of tested samples are shown, with colors indicating the clinical department that ordered each test. **C.** Age distribution of study participants across the five study-period categories.

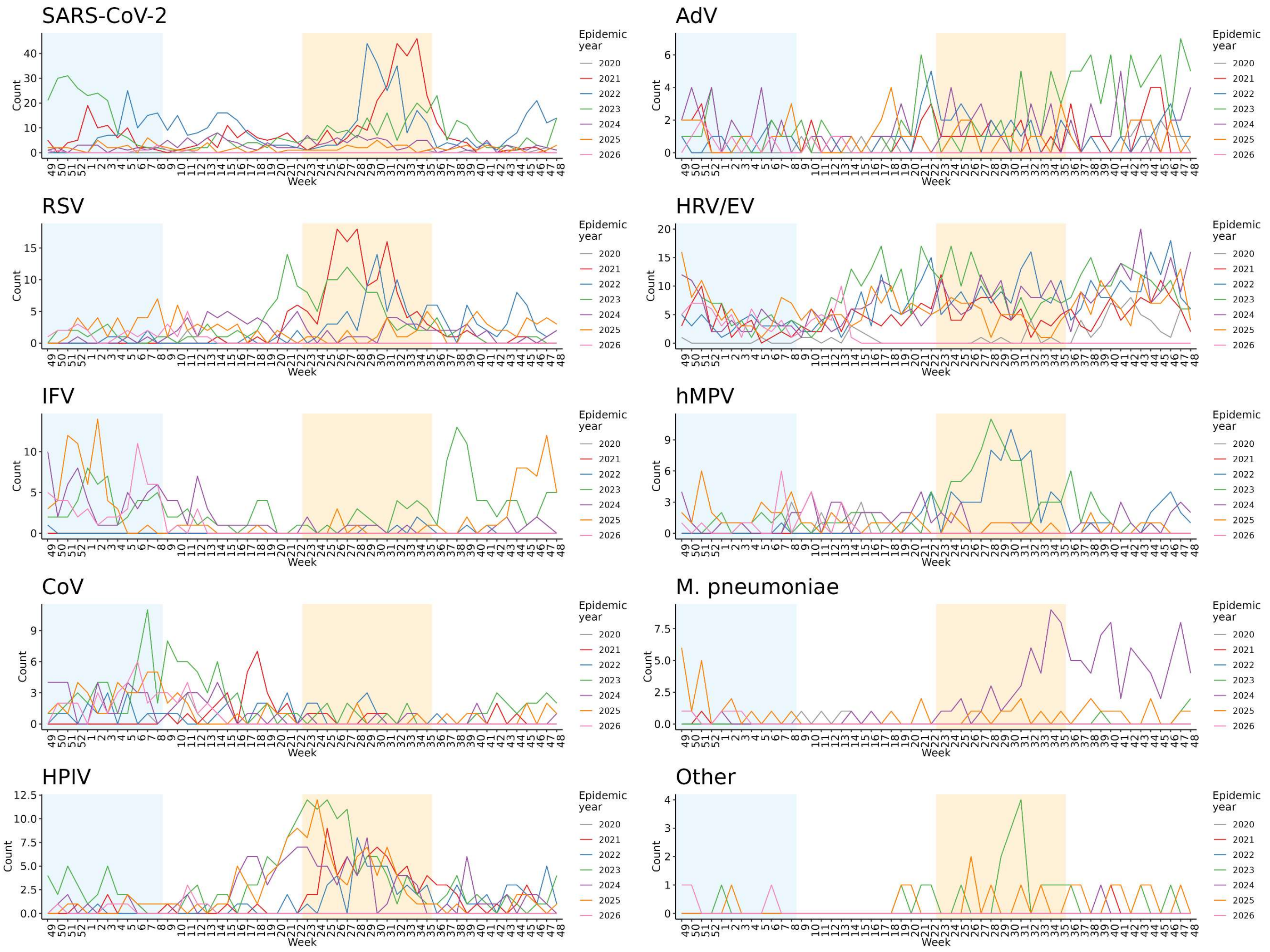


**Supplementary Figure 2. Seasonal trends in respiratory pathogens during each epidemic year.** Weekly counts of detected pathogens are shown for each respiratory pathogen. The x-axis shows week numbers, beginning in winter and ending in autumn. Colored lines represent epidemic years (2020−2026). Blue and orange backgrounds indicate winter and summer, respectively.

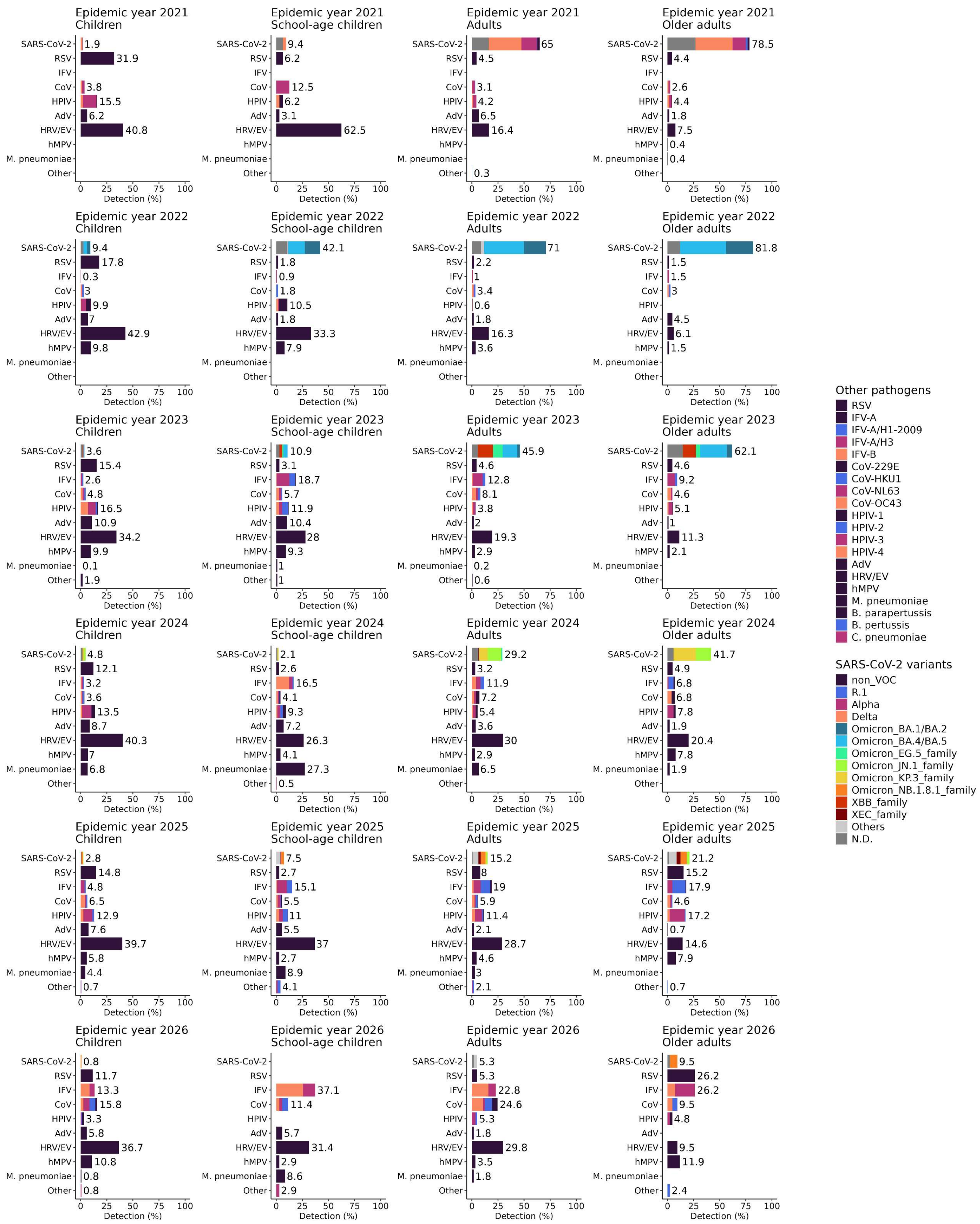


**Supplementary Figure 3. Compositions of respiratory pathogens by age group and epidemic year.** The bar lengths and the numbers shown behind them indicate the proportion of each respiratory pathogen, calculated as the number of positive detections for each pathogen divided by the total number of positive detections within each age group and epidemic year. Colors indicate individual species, lineages, or subtypes, as appropriate. N.D.: not determined.

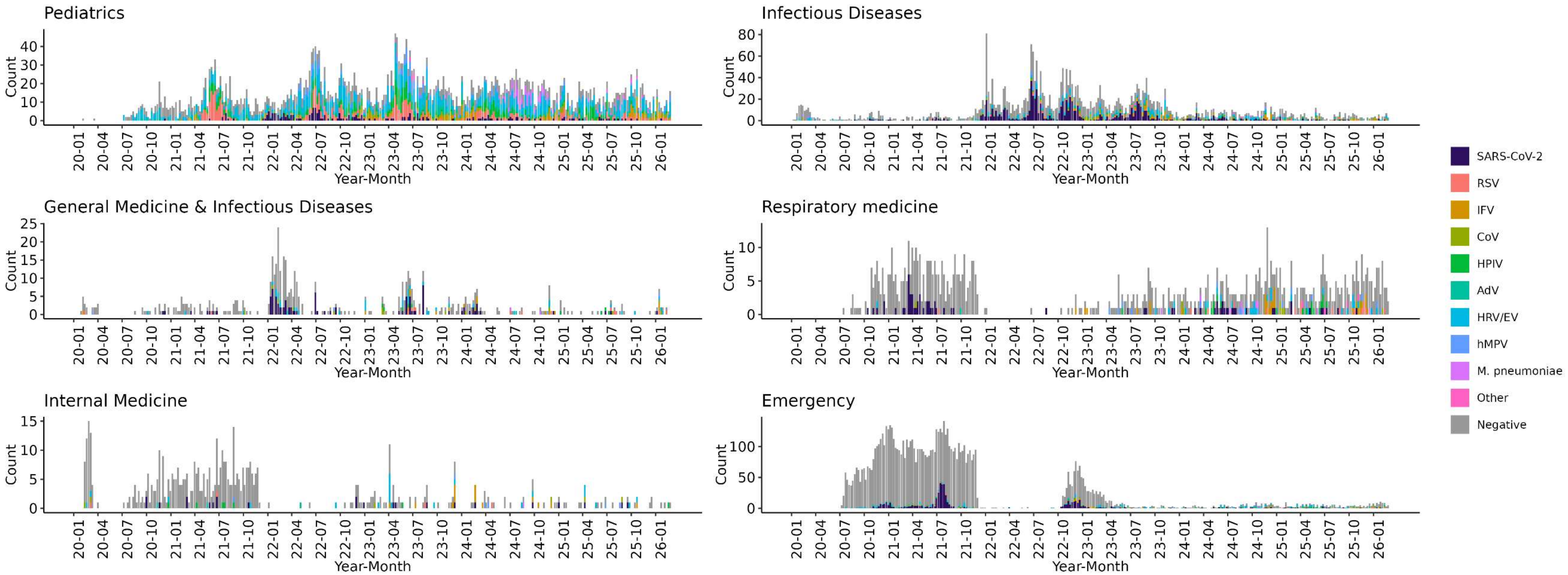


**Supplementary Figure 4. Distribution of respiratory pathogens by ordering clinical department.** Weekly counts of all tested samples are shown by the clinical department that ordered each test. Only departments that ordered more than 500 tests and had a positivity rate exceeding 10% were included. The y-axis shows the number of test results, with multiple pathogen detections from a single sample counted separately; colors indicate the detected respiratory pathogens, and gray indicates samples in which no pathogen was detected. The x-axis shows the year and month of sample collection.

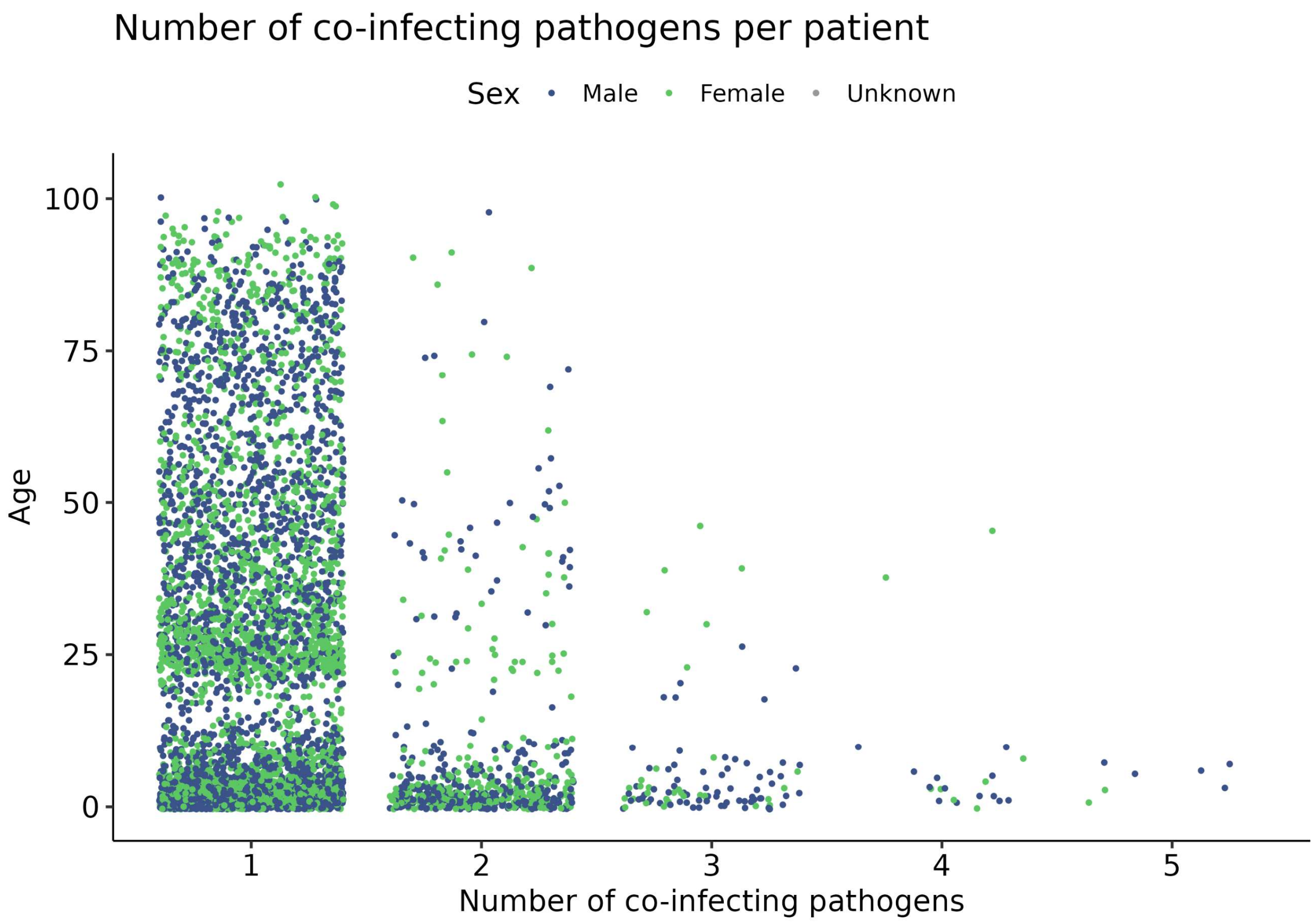


**Supplementary Figure 5.** Distribution of samples with multiple pathogen detections.
Patient age is shown according to the number of respiratory pathogens detected per sample. The x-axis shows the number of pathogens detected (1–5), and the y-axis shows patient age. Blue and green dots indicate male and female patients, respectively.

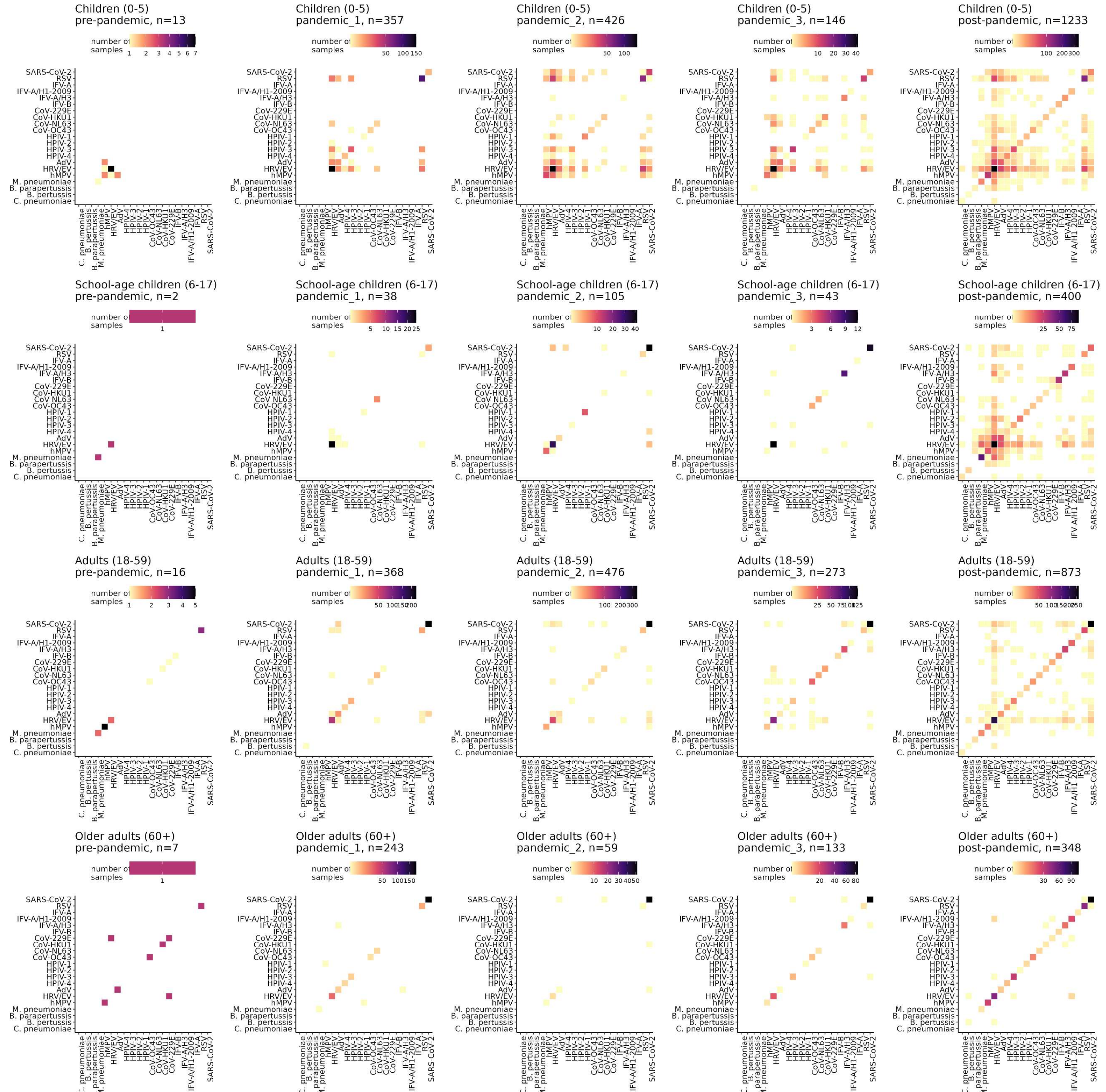


**Supplementary Figure 6. Pairwise co-detection patterns among respiratory pathogens by age group and study period.** The heatmaps illustrate pairwise co-detection patterns among respiratory pathogens. Each cell represents the number of samples in which the corresponding pathogen pair was detected, with diagonal cells representing single-pathogen detection frequencies. The analyses were restricted to samples with at least one detected pathogen. Color intensity indicates the number of samples, with lower and higher values represented by yellow and dark purple, respectively.

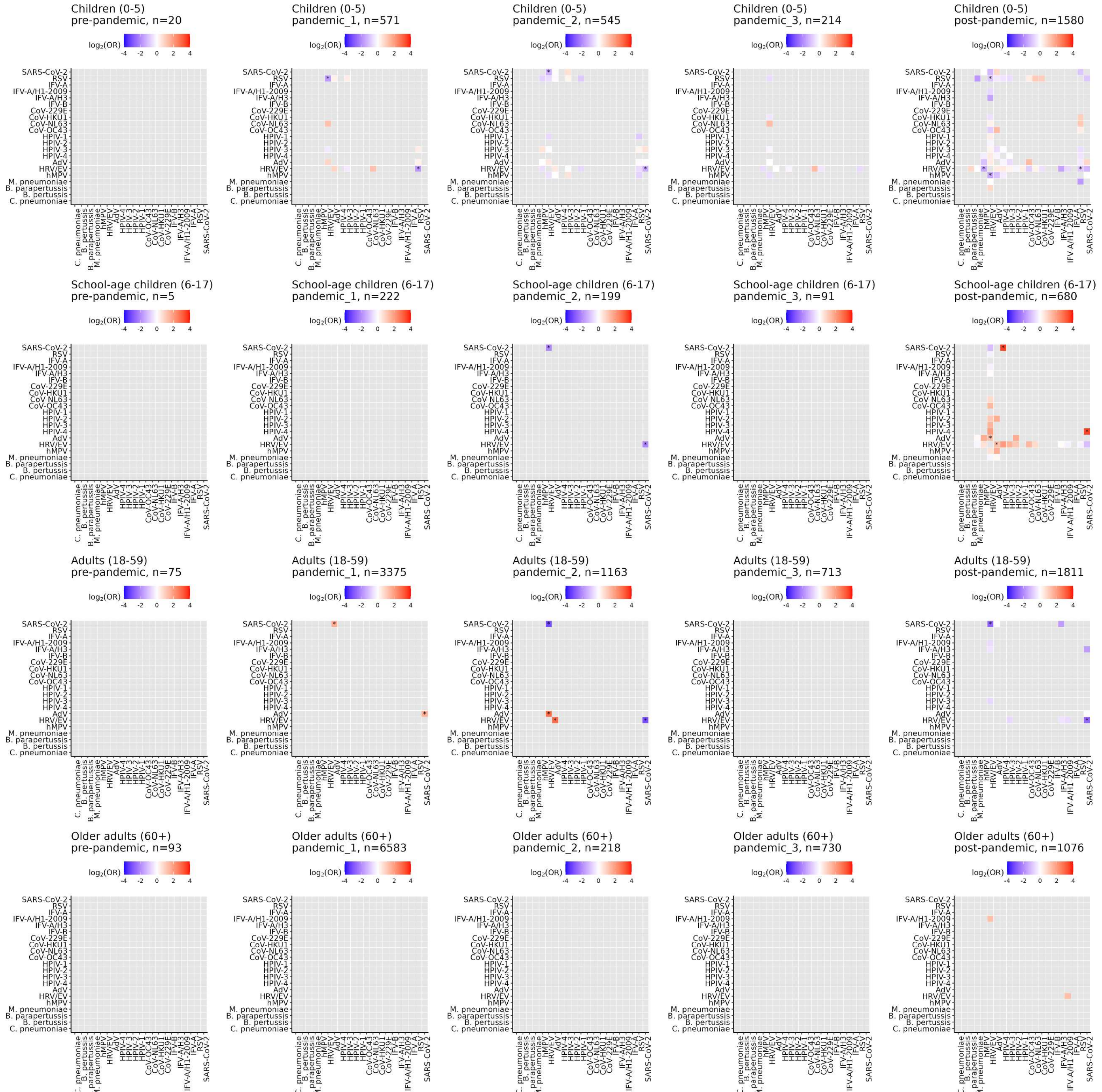


**Supplementary Figure 7. Association patterns among respiratory pathogens by age group and study period.** The heatmap shows statistical pairwise associations between pathogen pairs based on log2-transformed odds ratios (ORs). The analyses were performed on all tested samples, including those with no detected pathogen. Red and blue indicate positive and negative associations, respectively, and color intensity represents the strength of the association. Significant associations after false discovery rate (FDR) correction are indicated by an asterisk.

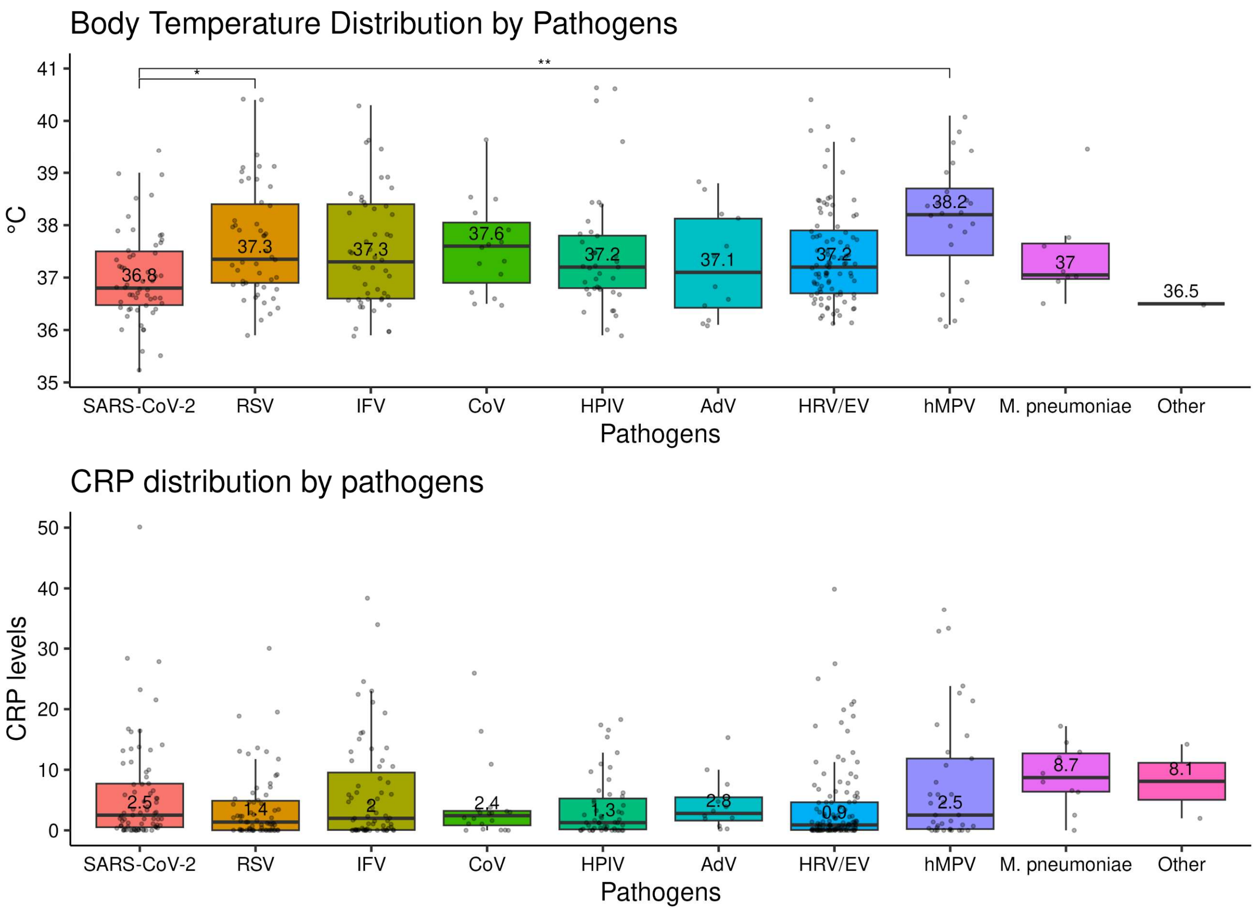


**Supplementary Figure 8. Associations between respiratory pathogen detection and clinical characteristics.** To exclude the influence of co-detection, only test records in which a single pathogen was detected were included. Furthermore, only records collected during the post-pandemic period, when symptomatic cases were more prevalent, were included. This resulted in a total analytical dataset of n=2,340. Of these records, body temperature data were available for n=344 and CRP levels for n=459. Groups were compared using the Kruskal-Wallis test, followed by Dunn's post hoc test with Benjamini-Hochberg adjustment (adjusted P values: * <0.05 and ** <0.01). Boxplots show the distributions of body temperature **(A)** and C-reactive protein (CRP) levels **(B)** by respiratory pathogen. The center lines and boxes indicate the median and interquartile range (IQR), respectively. Body temperature was significantly lower in the SARS-CoV-2 group than in the RSV and hMPV groups (p=0.028 and p=0.0019, respectively). Because patients with SARS-CoV-2 infection were older than those infected with other pathogens, we fitted a linear regression model adjusted for age, with SARS-CoV-2 as the reference group. The differences remained significant after adjustment for age, and IFV was also associated with a significantly higher body temperature than SARS-CoV-2 in the adjusted analysis. CRP is an acute-phase protein that serves as an early marker of inflammation or infection. Patients with bacterial infections have been reported to have higher CRP levels than those with viral infections [30]. In our data, CRP levels were higher in patients with *M. pneumoniae* and other bacterial and atypical bacterial infections; however, Dunn's post hoc test showed no significant differences. In contrast, in a linear regression model adjusted for age, with *M. pneumoniae* as the reference group, significant differences in CRP levels were observed for the RSV, HPIV, and HRV/EV groups (p=0.015, p=0.0094, and p=0.049, respectively). The "Other" category includes bacterial and atypical bacterial pathogens.

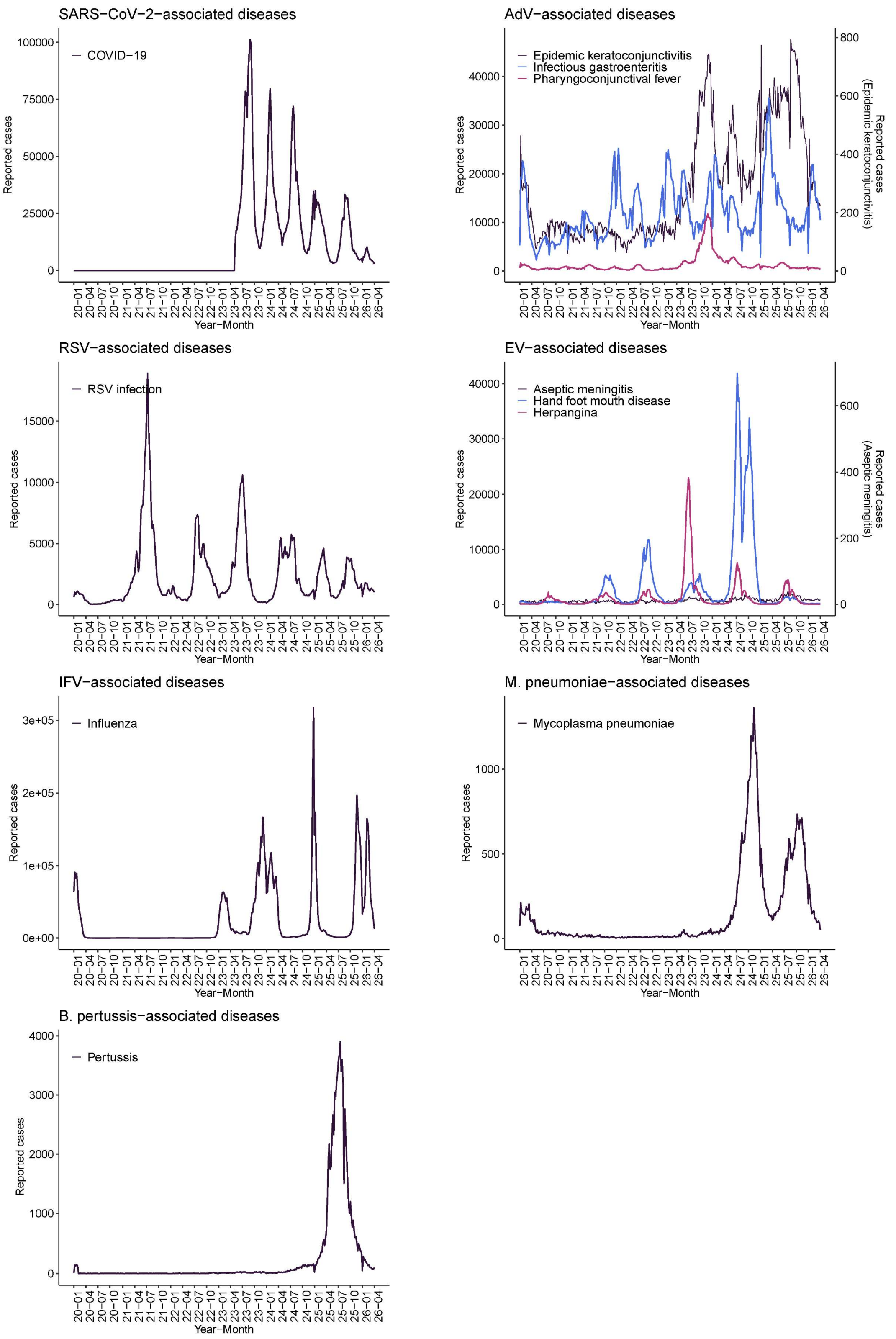


**Supplementary Figure 9. Weekly epidemiological trends of respiratory pathogen-associated diseases in Japan.** Data were obtained from the Infectious Diseases Weekly Report (IDWR) (https://id-info.jihs.go.jp/en/surveillance/idwr/rapid/index.html). Case counts were based on sentinel surveillance for all diseases except pertussis, which was based on all-case surveillance. Weekly case counts were plotted, with the x-axis labeled as Year-Month. COVID-19 case data were available from week 19 of 2023.

#### Supplementary Table 1. Age distributions by study period

| Age group | Pre-pandemic (tested n=193) | | | Pandemic 1 (tested n=10,754) | | | Pandemic 2 (tested n=2,125) | | | Pandemic 3 (tested n=1,748) | | | Post-pandemic (tested n=5,148) | | |
|---|---|---|---|---|---|---|---|---|---|---|---|---|---|---|---|
| | Total | Positive | Proportion | Total | Positive | Proportion | Total | Positive | Proportion | Total | Positive | Proportion | Total | Positive | Proportion |
| Children (0-5) | 20 | 13 | 65.0 | 571 | 357 | 62.5 | 545 | 426 | 78.2 | 214 | 146 | 68.2 | 1,580 | 1,233 | 78 |
| School-age children (6-17) | 5 | 2 | 40.0 | 222 | 38 | 17.1 | 199 | 105 | 52.8 | 91 | 43 | 47.3 | 680 | 400 | 58.8 |
| Adults (18-59) | 75 | 16 | 21.3 | 3,375 | 368 | 10.9 | 1,163 | 476 | 40.9 | 713 | 273 | 38.3 | 1,811 | 873 | 48.2 |
| Older adults (60+) | 93 | 7 | 7.5 | 6,583 | 243 | 3.7 | 218 | 59 | 27.1 | 730 | 133 | 18.2 | 1,076 | 348 | 32.3 |
| Unknown | | | | 3 | 0 | - | | | | | | | 1 | 1 | - |

#### Supplementary Table 2. Pathogen distributions by study period

| Pathogens | All (tested n=19,968) | | Pre-pandemic (tested n=193) | | Pandemic_1 (tested n=10,754) | | Pandemic_2 (tested n=2,125) | | Pandemic_3 (tested n=1,748) | | Post-pandemic (tested n=5,148) | |
|---|---|---|---|---|---|---|---|---|---|---|---|---|
| | Positive | Proportion | Positive | Proportion | Positive | Proportion | Positive | Proportion | Positive | Proportion | Positive | Proportion |
| SARS-CoV-2 | 1,662 | 8.3% | 0 | 0.0% | 428 | 3.98% | 509 | 24.0% | 241 | 13.8% | 484 | 9.4% |
| RSV | 652 | 3.3% | 4 | 2.1% | 147 | 1.37% | 116 | 5.5% | 28 | 1.6% | 357 | 6.9% |
| IFV | 426 | 2.1% | 1 | 0.5% | 1 | 0.01% | 8 | 0.4% | 64 | 3.7% | 352 | 6.8% |
| CoV | 340 | 1.7% | 7 | 3.6% | 37 | 0.34% | 36 | 1.7% | 78 | 4.5% | 182 | 3.5% |
| HPIV | 576 | 2.9% | 0 | 0.0% | 86 | 0.80% | 70 | 3.3% | 46 | 2.6% | 374 | 7.3% |
| AdV | 366 | 1.8% | 3 | 1.6% | 63 | 0.59% | 52 | 2.4% | 23 | 1.3% | 225 | 4.4% |
| HRV/EV | 1,918 | 9.6% | 12 | 6.2% | 324 | 3.01% | 351 | 16.5% | 157 | 9.0% | 1,074 | 20.9% |
| hMPV | 347 | 1.7% | 11 | 5.7% | 1 | 0.01% | 84 | 4.0% | 23 | 1.3% | 228 | 4.4% |
| M. pneumoniae | 164 | 0.8% | 4 | 2.1% | 1 | 0.01% | 0 | 0.0% | 0 | 0.0% | 159 | 3.1% |
| Bordetella spp. C. pneumoniae | 40 | 0.2% | 0 | 0.0% | 1 | 0.01% | 0 | 0.0% | 1 | 0.1% | 38 | 0.7% |
